# Seropositivity of HBV & HCV among the Family Members of Chronic Viral Hepatitis Patients

**DOI:** 10.1101/2021.05.12.21257115

**Authors:** Abu Bakar Siddique, Mohammad Nezam Uddin, Kajal Kanti Das, Rajat Sanker Roy Biswas

## Abstract

**Introduction:** Hepatitis B virus (HBV) and hepatitis C virus (HCV) infections are the global public health challenge. Family members of the HBV and HCV infected patients have a higher risk of exposure to many blood-borne diseases including Hepatitis B, and Hepatitis C viral infections as well. So the objectives of the present study was to evaluate seropositivity of HBV and HCV among family members of. chronic viral hepatitis patients.

**Methods:** This study was conducted 80 family members of 50 patients with chronic HBV and HCV infection. After ethical clearance and written consent, 1^st^ degree family members of those chronically infected hepatitis patients were explained about the objectives of the study and a standard questionnaire were introduced and recorded. Venous blood samples was taken from every participant with universal precautions and was tested by Enzyme Linked Immunosorbent assay (ELISA) for Hepatitis B surface antigen, anti HCV. Data analysis was done later by SPSS.

**Results:** Eighty family members of 50 HBV and HCV positive cases where screened where HBV infection(45;90%) was more then HCV infection(5;90%). Male were 45(56.3%) and female were 35 (43.8%) male to male ratio 1.28:1. Total 23 (28.8%) subjects were vaccinated against HBV. One subject had history of blood transfusion and dental procedure each, extramarital sex activity was found in 2 (2.5%), and sleeping in the same room was found in 31(38.6%) subjects. History analysis revealed, 3(3.8%) were positive for HBV, 29 (36.3%) were found negative and 48(60%) were unaware of their status; again 77 (96.3%) are unaware of their anti HCV status. HBsAg and Anti HCV screening status of the relatives revealed HBsAg and anti HCV was found positive in 4 (5%) and 1 (1.3%) cases respectively.

**Conclusion:** Relatives of the HBV and HCV infected patients are also at risk. So they should take proper preventive measures and should be vaccinated.

## Introduction

Globally there are about 360 million chronic carriers of HBV and over one million people die each year as a result of acute fulminant liver disease or HBV induced cirrhosis and liver cancer [1].The burden of HBV infection is highest in the developing world particularly Asia and sub-Saharan Africa [2–4]. In the Middle East and Indian sub-continent, HBV infection is of intermediate endemicity with chronic HBV carriage rate of 2-5% among general population [5].

One of the most substantial problems in public health is hepatitis C virus (HCV) infection, which affects approximately 1%-5% of the world population and occurs in all countries. Epidemiological information on HCV is essential for strategic prevention of chronic hepatitis, liver cirrhosis and cancer. The rate of HCV infection differs in particular countries. The prevalence in developed countries amounts to 0.2%-2.2%, while in developing countries it reaches 7%. In some regions or in risk groups the rate of occurrence may be as high as 30%-90%[6].

HBV and HCV infection is acquired mainly parenterally by transfusion of infected blood, rupture of the continuity of skin or mucous membrane, infected medical equipment despite strict hygienic control, intravenous drug abuse, hemodialysis or organ transplantation. HCV infection is an important cause of post-transfusion hepatitis. Transmission through sexual contacts has been implicated, although this may be a rather inefficient mode of transmission. HCV has also been detected in persons in whom no clear risk factor has been defined, and these cases constitute about 40%-45% of HCV infections[7]. Mother-to-infant transmission has also been demonstrated [6] but the possibility of other transmission routes has not been thoroughly explored. With the use of RT-PCR or bDNA techniques, HCV RNA has been detected in many systemic fluids other than in blood, including peritoneal effusion, seminal and vaginal secretion, urine, feces and typhoid secretion. At least 20% of hepatitis C patients develop cirrhosis with the associated risk of developing hepatocellular carcinoma (HCC)[7].

To our knowledge little information is available about HBV and HCV infections due to contacts of patients infected with HCV in Bangladesh. Because HBV and HCV may be transmitted by the non-parenteral routes such as sexual and non-sexual household contacts, this study was undertaken to investigate whether intrafamilial transmission occurs *via* the usual contacts between patients and their household members who are unaware of the potential infectious state of the patients, to determine the prevalence of antibody to HCV in the contacts of HCV positive cases (index patients) and to evaluate the potential risk factors associated with intrafamilial transmission of HBV and HCV.

In Bangladesh, there is paucity of information on the prevalence of HBV and HCV infections among the family members and majority of the previous studies were conducted in selected group of people such as blood donors, drug addicts, commercial sex workers (CSWs) or hospitalized patients [8]. However, a recent report showed 5.5% HBsAg positivity among the general population living in Savar, a semi-urban area on the outskirts of Dhaka [9,10]. As there is little information on HBV and HCV among family members of HCV and HBV infected patients, we decide to estimate HBV and HCV infection status among those family members in a tertiary care hospital.

## Methods

It was a cross sectional descriptive study done in the Departments of Medicine of a tertiary care hospital, Chittagong, Bangladesh during a study period of one year. Sample size was 80 and sampling technique was purposive type. 1^st^ degree family members those who were attending or staying with chronic HBV and HCV infected patients admitted in the hospital were taken as study population. If wife was infected husband and children were screened, if husband infected wife and children were screened, if unmarried parents, brothers and sisters were screened. After written consent, all the family members were counseled in one-to-one manner and explained about the objective of the study. Each personal interview was carried out in full respect of confidentiality. All family members were provided with the results of their serological tests and any further information they wished to have. Face-to-face interviews was performed by the researcher himself. A structured questionnaire was used to record sociodemographic and prior symptomatic hepatitis, HBV vaccination status, occupational and non-occupational risk factors. Work-related risk factors include questions about occupation, instances of blood exposure. Non-occupational risk factors include questions about e.g. blood transfusion, dialysis and surgery and individual lifestyle risks. Using an aseptic technique, about 5 ml of venous blood was drawn from each participant and immediately put in a vacutainers containing a clot activator. The vacutainers were labeled indicating the serial number and date of sample collection. Blood samples was then taken to the laboratory for separation of sera. Then eera was stored at −20°C awaiting investigations. We used Enzyme Linked Immunosorbent Assay (ELISA), (Stat Fax-3200, USA, and reagent from Enzo, USA) to carry out HBsAg, and Anti HCV tests. Results were determined spectrophotometrically and interpretation of test results was done as per manufacturer’s guidelines. An informed consent was sought from the patient. Patient’s data was recorded on pre designed case record form (CRF). All the parameters was analyzed with appropriate statistical test if needed. Statistical analysis was performed with the SPSS-20 software package(IBM SPSS, Armonk, NY, USA)

## RESULTS

We have screened 80 family members of total 50 HBV and HCV positive cases where HBV infection(45, 90%) was more then HCV infection(5, 90%)(Table I). Table II showing 45(56.3%) were male and rest 35(43.8%) were female. Male to female ratio was 1.28:1. Among all 23(28.8%) were found to be vaccinated against HBV (Table III). Table IV showing different risk behaviors of the study subject where history of blood transfusion and dental procedure was found in one subject each, extramarital sex activity was found in 2(2.5%), and sleeping in the same room was found in 31(38.6%) subjects. In history analysis we found, 3(3.8%) were known to be positive for HBV, 29(36.3%) were found negative and 48(60%) were unaware of their status; again 3(3.8%) were known to be negative for anti HCV and 77(96.3%) are unaware of their anti HCV status. (Table V). Table VI showing HBsAg and Anti HCV screening status of the relatives of the infected patients where HBsAg was found positive in 4(5%) cases and Anti HCV was found positive in one case.

**Table I:** Pattern of infected patients recruited (n=50)

| Infection | Frequency | Percent |
| --- | --- | --- |
| HBV | 45 | 90.0 |
| HCV | 5 | 10.0 |
| Total | 50 | 100.0 |

**Table II:** Gender of the study subjects screened(n=80)

|  | Frequency | Percent |
| --- | --- | --- |
| Female | 45 | 56.3 |
| Male | 35 | 43.8 |
| Total | 80 | 100.0 |

**Table III:** HBV vaccination status(n=80)

|  | Frequency | Percent |
| --- | --- | --- |
| Yes | 23 | 28.8 |
| No | 57 | 71.3 |
| Total | 80 | 100.0 |

**Table IV:**
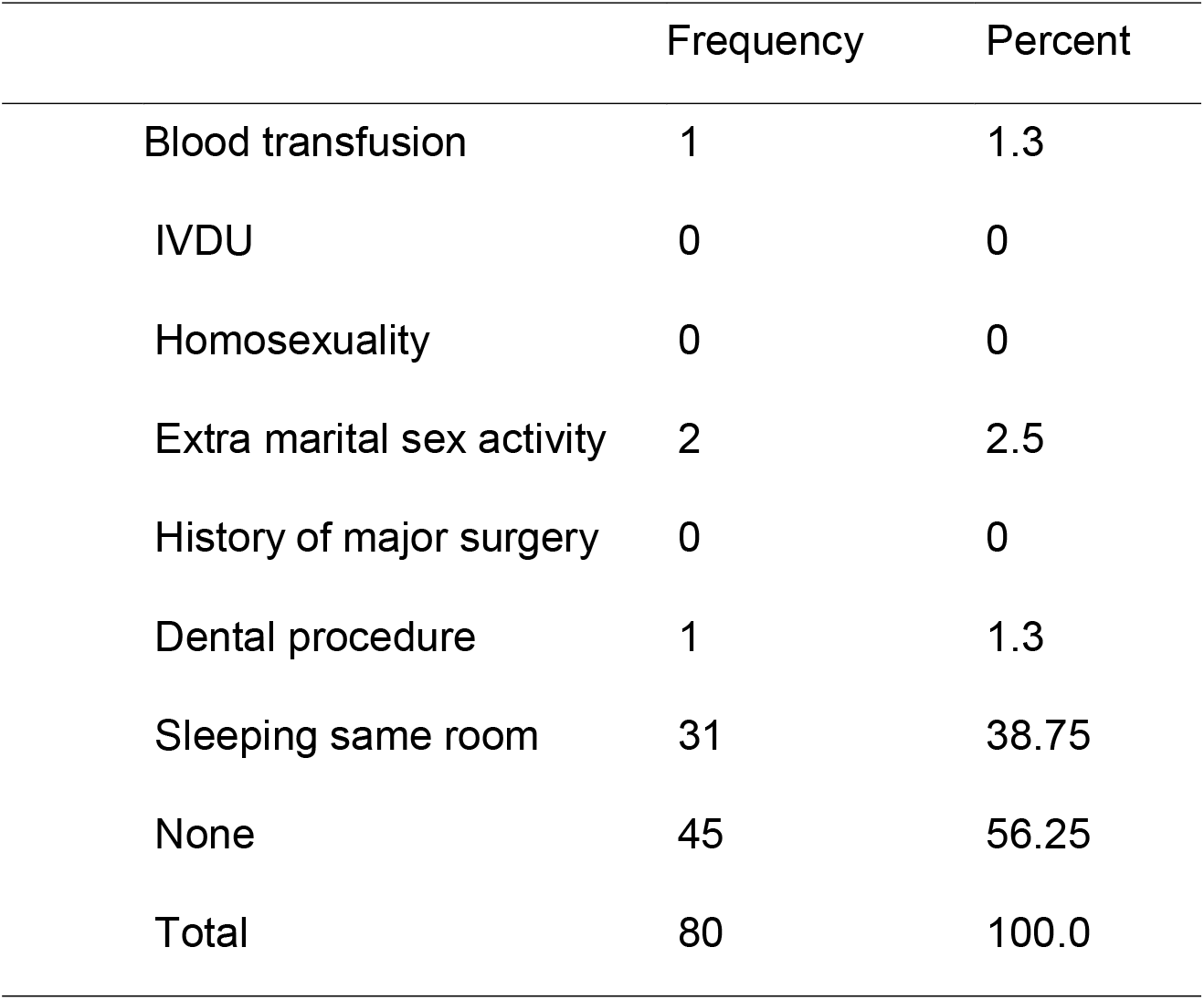
Risk behaviors(n=80)

|  | Frequency | Percent |
| --- | --- | --- |
| Blood transfusion | 1 | 1.3 |
| IVDU | 0 | 0 |
| Homosexuality | 0 | 0 |
| Extra marital sex activity | 2 | 2.5 |
| History of major surgery | 0 | 0 |
| Dental procedure | 1 | 1.3 |
| Sleeping same room | 31 | 38.75 |
| None | 45 | 56.25 |
| Total | 80 | 100.0 |

**Table V:** Past history of HBV and HCV status(n=80)

|  |  | Frequency | Percent |
| --- | --- | --- | --- |
| HBV | Positive | 3 | 3.8 |
|  | Negative | 29 | 36.3 |
|  | Not known | 48 | 60.0 |
|  | Total | 80 | 100.0 |
|  |  | Frequency | Percent |
| HCV | Negative | 3 | 3.8 |
|  | Not known | 77 | 96.3 |
|  | Total | 80 | 100.0 |

**Table VI:** Investigation report of HBsAg and anti HC(n-80)V

|  |  | Frequency | Percent |
| --- | --- | --- | --- |
| HBs Ag | Positive | 4 | 5.0 |
|  | Negative | 76 | 95.0 |
| Anti HCV | Positive | 1 | 1.3 |
|  | Negative | 79 | 78.7 |

## Discussion

This study was conducted in a tertiary care hospital in Bangladesh where 80 relatives of total 50 HBV or HCV infected persons were included as they were supposed to at a greater risk of various blood-borne infections including hepatitis B and hepatitis C.

Among 50 selected patients of HBV or HCV positive cases 45(90%) cases infected with hepatitis B and 5(10%) were infected with hepatitis C. In Bangladesh infection of HBV is about 6.5% and HCV is 1% in general population.[11]. So here in our findings it is also reflected as there are more HBV positive cases recruited.

Male family members were more in the study populations which was 45(56.3%) and rest 35(43.8%) were female. Male to female ratio was 1.28:1. In this study more male were recruited as male commonly come forward in dealing health related matters of their relatives in our country.

In our study, we found that out of 80 subjects 23(28.8%) were found to be vaccinated against HBV. The proportion of vaccinations among relatives varies. The reason of such differences of vaccination coverage can be explained on the basis of their educational and awareness status. The percentage of vaccinated among the community is much higher, as estimated by WHO in the South-East Asian Region (SEAR). According to the World Health Organization (WHO) estimates, hepatitis B vaccination coverage among relatives and health care workers varies from 18% in Africa to 77% in Australia and New Zealand.[12]

When history was analyzed regarding the HBV and HCV only 3(3.8%) were known to be positive for HBV and none were aware of their HCV status. This is a fact in Bangladesh that though there are known positive cases of HBV and HCV in the family they are reluctant to do the test for screening.

Regarding screening of HBsAg and Anti HCV status of the 80 relatives of the known infected patients where HBsAg was found positive in 4(5%) cases and Anti HCV was found positive in one case. A study conducted in Dhaka, Bangladesh by Ashraf *et al*.[13] found that the HBsAg prevalence of 6.5% among the study population was within the range of 2-7%, as reported by previous studies from selective populations of Dhaka, Bangladesh: 3% among healthy adults and children and 3.5% among pregnant women; however, a much lower rate (0.8%) was observed among schoolchildren. Among health care workers it was found 8% in Bangladesh[11]. Again HCV infection is 1% in general and in our finding among the relatives it is similar. So our study findings suggested that the relatives of these groups of patients infected with HBV or HCV have a similar chances to be infected as the general population of Bangladesh.

## Conclusion

So we can conclude that relatives of the HBV and HCV infected patients are also at risk but not exceeding the risk for general population of Bangladesh. So they should take proper preventive measures.

## Data Availability

Data is available

## Limitations

Single center study and small sample size

## Recommendations

Regular screening of relatives

Proper preventive measures like vaccination should be taken

Large scale multicenter trial to see the national scenario

## Conflict of interest

None

## Notes

### Competing Interest Statement

The authors have declared no competing interest.

### Author Declarations

ERB clearance was taken from ERB CMOSHMC

